# Reduction of pro-inflammatory effector functions through remodeling of fatty acid metabolism in CD8^+^ T-cells from Rheumatoid Arthritis patients

**DOI:** 10.1101/2022.09.22.22280236

**Authors:** Franziska V. Kraus, Simon Keck, Karel D. Klika, Jürgen Graf, Rui A. Carvalho, Hanns-Martin Lorenz, M Margarida Souto-Carneiro

## Abstract

**Objectives:** RA CD8^+^ T-cells (CD8) maintain their effector pro-inflammatory phenotype by changing their metabolism towards aerobic glycolysis. However, their massive energetic and biosynthetic needs may require additional substrates to furnish this high demand. Since systemic alterations in lipid metabolism have been reported in RA patients, we explored the role of fatty acid (FA) metabolism in CD8 to identify potential targets to curb their pro-inflammatory potential.

**Methods:** The expression of FA metabolism-related genes was analyzed for total and CD8-subsets from RA patients and healthy controls (CNT). Peripheral-blood CD8 were isolated from RA, PsA, SpA patients under different therapies (DMARD, biologicals, JAK-inhibitors) and CNT and were TCR-stimulated with or without FA metabolism inhibitors. We quantified the expression of the main FA transporters, lipid uptake, intracellular content of (un-)saturated FA, cytokine production, activation, proliferation, and capacity to inhibit tumor cell growth.

**Results:** The CD8 gene expression profile of FA metabolism-related genes was significantly different between untreated RA patients and CNT. RA patients with a good clinical response after 6 months MTX therapy significantly increased the expression of FA metabolism-related genes. Cell-surface expression of FA transporters FABP4 and GPR84 and FA-uptake was higher in effector and memory CD8 of RA patients than for CNT. *In vitro* blockade of FA metabolism significantly impaired CD8 effector functions.

**Conclusions:** RA CD8 present an altered FA-metabolism which can be potential therapeutic targets to control their pro-inflammatory profile, especially by targeting the transport and oxidation of free FAs which are abundant in the serum and synovial fluid of patients.

## Introduction

The treatment of chronic inflammatory autoimmunopathies such as rheumatoid arthritis (RA) has greatly improved with the introduction of cytokine blockers, cell-targeted therapeutic antibodies, and Janus kinase (JAK) inhibitors. However, in contrast to psoriasis, complete remission of arthritis in the majority of patients over a prolonged treatment period is still an exception, indicating that we might be missing an important molecular component in the initiation and/or chronic propagation of the inflammatory response leading to chronic arthritis as exemplified by RA.

Nevertheless, the interest in fatty acid (FA) metabolism involvement in RA dates back to the 1960s where initial studies revealed 40 % -60 % more phospholipids and cholesterol in the synovial fluid of RA patients compared to the respective serum samples, and, moreover, the increase of synovial FAs correlated with the increase of synovial protein concentration [1]. Later, it was found that cholesterol-regulating therapies and low-fat diets could improve the symptoms of autoimmune diseases and the T cell-dependent auto-antibody response [reviewed in 2], thus further underlining the role of lipid metabolism in chronic inflammation and autoimmunity. On this basis, we wanted to characterize lipid metabolism in the lymphocytes of RA patients and to investigate the influence of immunosuppressive therapy. Upon activation, T cells upregulate both glycolysis and FA and cholesterol synthesis to provide enough energy to meet biosynthetic demands and clonal expansion [3], and then return to oxidative phosphorylation (OXPHOS) and FA oxidation (FAO) when the effector functions are no longer required. However, in chronic (auto)inflammation, T cells maintain a permanent activated profile which is sustained by an aberrant metabolic profile [4]. Studies have shown that RA CD4^+^ T cells (CD4) have reduced mitochondrial activity which prevent the oxidative usage of FAs, leading to an accumulation of intracellular lipid droplets and changes in cellular FA metabolism that are linked to increased pro-inflammatory effector functions but can be abrogated by the FA synthase inhibitor C57 [5]. While CD4 FA metabolism has proven to be a promising research target in RA, CD8^+^ T cell (CD8) lipid metabolism in RA remains largely unexplored. In a previous study we presented evidence that CD8 in RA reveal an altered, hyper-glycolytic metabolic profile which is accompanied by an increased glucose-derived synthesis of sphingophospholipids [4]. Thus it is likely that further studies of FA metabolism -not only of CD4 but also of CD8 - will reveal new insights into the immunometabolic mechanisms that maintain chronic inflammation in RA.

Upon CD8 T cell receptor (TCR) activation, metabolic reprogramming is initiated and mTORC1 mediated SREBP signaling upregulates cholesterol and FA synthesis-related enzymes like ACC, FASN, and HMGCR which are proven to be necessary processes for clonal expansion [6–9] and the acquisition of effector functions. Moreover, it has been shown that the synthesis of carnitines as well as the degradation of palmitate, which are needed for FA oxidation (FAO), are decreased in stimulated T-cells and *de novo* FA synthesis (FAS) is initiated to increase cellular biomass, in a myc dependent manner[10]. Indeed, human RA CD8 contain elevated amounts of neutral lipids when compared to healthy CD8 [4]. While tissue resident CD8 memory cells seem to satisfy their FA metabolism requirements by increased FA import and FAO, central memory CD8 fuel FAO with *de novo* synthesized FA and triglycerides [11,12].

In RA CD8, these differentiation-related demands for FA availability seem not only to be mediated by cell-intrinsic alterations in FA synthesis (FAS) or FAO pathway usage, but also by differential FA transporter expression [7]. The FA uptake in CD8 is mainly regulated by three families of cell surface transporters: CD36 [13,14], FA binding proteins/FA transporting proteins (FABP/FATP, especially FABP4 and FABP5) [11], and G-protein-coupled receptors (GPCR, especially GPR84) [15]. Accordingly, recent findings have shown augmented palmitate and long-chain FA uptake rates by effector CD8 [4,12].

The present study explores the importance of transporter-controlled FA uptake in CD8 in RA patients under different therapeutic regimens as well as the impact of FA metabolism modulation on CD8 cytotoxic capacities (Figure 1A).

**Figure 1:**
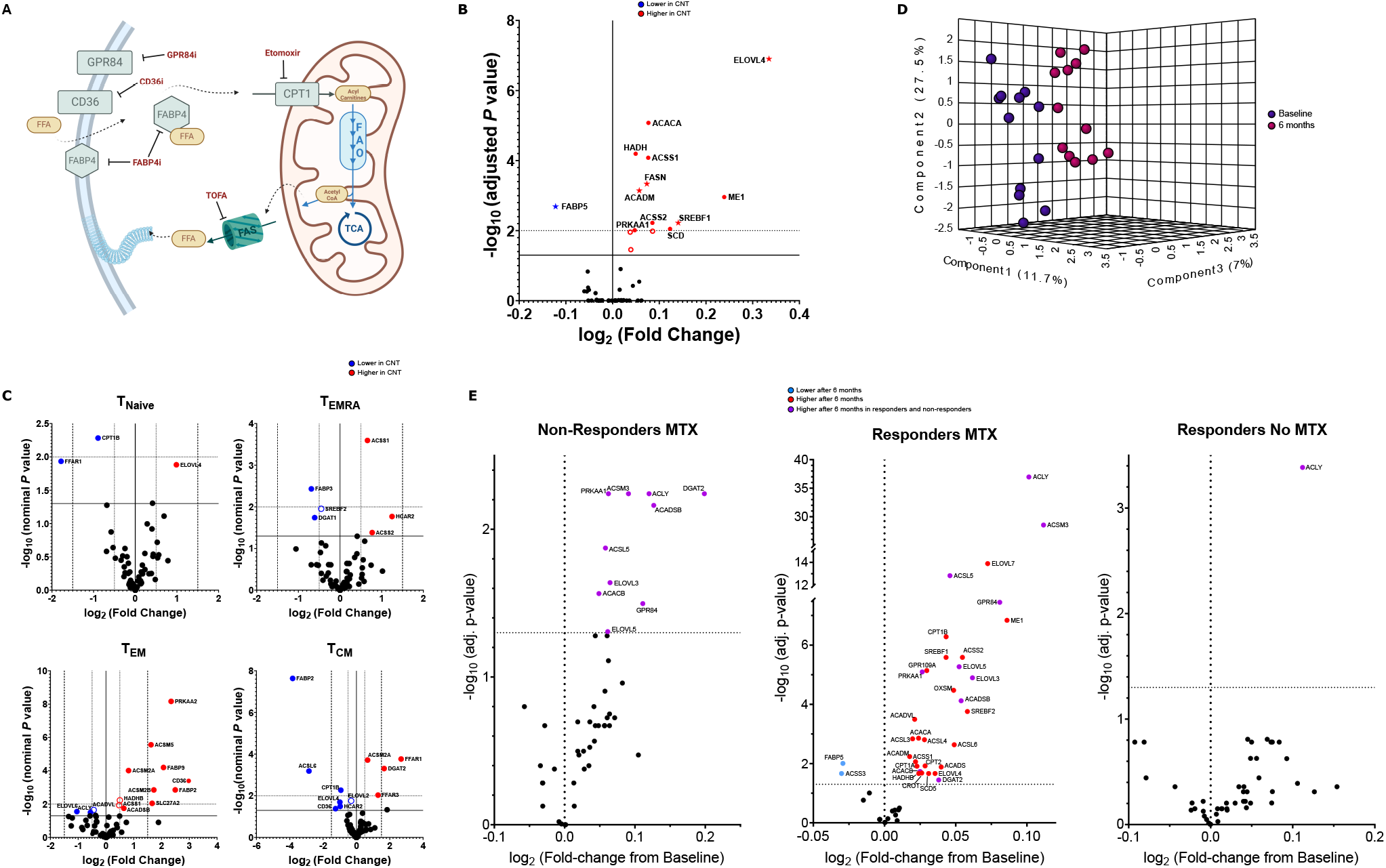
Gene expression of lipid metabolism related genes in CNT vs. RA CD8. **A** Schematic representation of the FA metabolism pathways in CD8, their major cell-surface transporters and the points-of-action of the inhibitors used in this study (created with BioRender.com). **B** Volcano plot of the gene expression differences for total CD8 between CNT and therapy naïve RA patients (CNT n=32; RA n=18). **C** Volcano plots of the gene expression differences in CD8 functional subsets between CNT and therapy naïve RA patients (n=7-10 per subsets per group). CNT vs. RA – blue = increased in RA; red = increased in CNT. **D** Paired PLS-DA with 3 principal components comparing CD8 from RA patients before and after 6 months of therapy. **E** Volcano plot of the gene expression differences for total CD8 from RA patients at baseline and after 6 months of therapy with either MTX (in combination with other DMARDs or alone) or other DMARD (no MTX) (MTX responders n=100; MTX non-responders n=13; no MTX responders n=10). FFA, free fatty acids; FAS, fatty acid synthesis; FAO, fatty acid oxidation; TCA, tricarboxylic acid cycle; T_NAIVE_, CD8 naïve subset; T_EMRA_, CD8 effector subset; T_EM_, CD8 effector memory subset; T_CM_, CD8 central memory subset.

### Patients, Material and Methods

A detailed description of the patient recruitment and selection process and of the experimental and statistical methodologies are described in detail in the accompanying supplementary methods. The clinical and demographic characteristics of the study cohort are summarized in Table 1.

**Table T1.** Clinical and demographic data of the study patients

|  | Healthy controls<br>(n=14) | RA<br>(n=31) | PsA<br>(n=26) | SpA<br>(n=21) |
| --- | --- | --- | --- | --- |
| **Female:Male** | 6:8 | 18:13 | 15:11 | 6:15 |
| **Age mean (min – max in years)** | 36.71 (23–58) | 59.52 (35–83) | 54.69 (36–71) | 50.86 (22–68) |
| **Disease duration mean (min – max in years)** | N.a. | 14.32 (0.5–44) | 9.6 (0.5–30) | 16.71 (1–45) |
| **DAS28-CRP mean (min – max in years)**<br>% active (DAS28 > 3.2)<br>% remission (DAS28 < 2.6) | N.a. | 2.25 (0.96–4.04)<br>19.4%<br>51.6% | 2.57 (0.97–6.48)<br>23.1%<br>53.8% | N.a. |
| **BASDAI mean (min – max)**<br>% active (BASDAI > 3.5)<br>% remission (BASDAI < 2.0) | N.a. | N.a. | N.a. | 3.15 (0.6–6)<br>33.34%<br>28.6% |
| **CRP mean ± SEM (mg/L)** | N.a. | 4.78±0.86 | 6.55±1.74 | 5.08±1.05 |
| **RF (positive:negative)** | N.a. | 24:7 | 0:26 | 0:21 |
| **Anti-CCP (positive:negative)** | N.a. | 28:3 | 0:25 | 0:21 |
| **HLA-B27 (positive:negative)** | N.a. | N.a. | 0:26 | 16:5 |
| **BMI mean** | 24.25 | 26.11 | 28.87 | 26.41 |
| **MTX**<br>Mean ±SEM (mg)<sup>a</sup><br>% from total | N.a. | 13.29±0.79<br>61.3% | 14.04±1.31<br>50% | 15<br>9.5% |
| **Glucocorticoid**<br>Mean ±SEM (mg/d)<sup>a</sup><br>% from total | N.a. | 3.73±0.65<br>32.3% | 6.4±1.23<br>34.6% | 5<br>9.5% |
| **Glucocorticoid + MTX**<br>% from total |  | 19.4% | 19.2% | 4.8% |
| **Leflunomid**<br>Mean ±SEM (mg)<sup>a</sup><br>% from total | N.a. | 16.9±1.32<br>25.8% | 17.5±2.5<br>7.7% | 10<br>9.5% |
| **COX-2 inhibitors**<br>Mean ±SEM (mg)<sup>a</sup><br>% from total | N.a. | 60±0<br>3.8% | 100±0<br>3.8% | 40.71±16.9<br>19% |
| **bDMARDs**<br>Anti-TNF (% from total)<br>Anti-IL-6R (% from total)<br>Anti-IL-17A (% from total)<br>Anti-IL-12/IL-23 (% from total)<br>JAK-blockade (% from total) | N.a. | 29.0%<br>6.5%<br>0%<br>0%<br>16.1% | 26.9%<br>0%<br>3.8%<br>7.7%<br>19.2% | 38.1%<br>0%<br>4.8%<br>9.5%<br>0% |

Legend: N.a., not available; SEM, standard error of the mean; DAS28, Disease Activity Score; BASDAI, Bath Ankylosing Spondylitis Disease Activity Index; CRP, C-reactive protein; RF, rheumatoid factor; anti-CCP, anti-cyclic citrullinated peptide antibodies; MTX, methotrexate. a) The average dosage of each medication (MTX, glucocorticoid, Leflunomid, COX-2 inhibitors) was calculated only for the patients taking that medication.

All participants gave their informed, written consent and the study was approved by the Institutional Ethics Committee of Heidelberg University (files S-096/2016 and S-969/2020).

## Results

### Expression analysis of lipid metabolism genes in RA CD8

To investigate FA and cholesterol metabolism and transport in the total CD8 pool from RA patients and healthy donors (CNT), we analyzed the differential gene expression before and after therapy initiation in addition to the related clinical and demographic data from the Gene Omnibus datasets GSE97475, GSE97948, and GSE97810 (cohort demographic and clinical characteristics have been described in [16]). To understand whether these differences were subset dependent, we analyzed the Gene Omnibus dataset GSE118829 in which the total CD8 pool of healthy donors and RA patients under different therapy regimens (none, MTX, IL-6R, or TNF blockade) were separated into functional subsets according to their CD45RA and CCR7 expression: naïve (CD45RA^+^CCR7^+^), effector (T_EMRA_) (CD45RA^+^CCR7^-^), effector memory (T_EM_) (CD45RA^-^CCR7^-^), and central memory (T_CM_) (CD45RA^-^ CCR7^+^) (clinical and demographic characteristics of this cohort are described in [17]). We also investigated how different therapies influenced lipid metabolism-related gene expression. Compared to CNT, CD8 from therapy-naïve RA significantly decreased the expression of several genes linked to lipid metabolism, whereas only *FABP5* was significantly overexpressed in RA (Figure 1B). Compared to untreated RA patients, we did not observe significant changes in the expression of several genes involved in FA metabolism, irrespective of whether the patients were treated with csDMARDs alone or in combination with glucocorticoids (GC) (Supplementary Figure 1A). The gene expression profiles were unique for the respective RA CD8 subsets (Figure 1C).

RA naïve CD8 showed a higher expression of FA transport-related genes (*CPT1B* and *FFAR1*), while CNT CD8 increased *ELOVL4*, a gene involved in FA elongation. In the effector compartment (T_EMRA_), both RA and CNT CD8 displayed an increased expression of genes involved in FA and cholesterol synthesis and FA transport (RA: *SREBF2, DGAT1*, and *FABP3*; CNT: *ACSS1, ACSS2*, and *CNTAR2*). In the T_EM_ population, CNT CD8 upregulated the expression of a variety of genes for FA transport and sensing (*PKAA2, FAPB2, FABP9, CD36*, and *SLC27A2*) as well as for acetyl-Co-A synthesis (*ACSS1, ACSM2A, ACSM2B*, and *ACSM5*), acetyl-Co-A breakdown (*ACADSB*), and FAO (HADHB). Similarly, in RA, T_EM_ genes involved in FAS (*ELOVL6* and *ACLY*), and also in FAO (*ACADVL*), were increased compared to CNT. The gene expression in the T_CM_ compartment resembled that of the T_EM_ compartment. Gene expression differences between CNT and RA in the T_EM_ and T_CM_ subsets were mostly related to FAS-, transport- and sensing-related genes. CNT CD8 T_EM_ and T_CM_ subsets increased the expression of *FFAR1, FFAR3, ACSM2A*, and *DGAT2* while in RA there was increased expression of *FABP2, CD36, CPT1B, CNTAR2, ACSL6, ELOVL2*, and *ELOVL4*. Paired partial least-squares differential analysis (PLS-DA) with three principal components of the subgroup that did not receive any type of therapy at baseline and after receiving DMARD for the first 6 months showed a significant separation of the two time points (accuracy: 87.5%; R^2^=0.935; Q^2^=0.535; Figure 1D; Supplementary Figure 1B). Next, we determined whether patients who responded to therapy after 6 months changed their FA metabolism-related gene expression profile. We observed an extensive alteration in gene expression in patients who responded to MTX therapy (combined with other DMARD or as monotherapy), but not in patients who responded to other therapeutic regimens, and only few changes were observed in those patients who did not respond to MTX therapy (Figure 1E).To validate the influence of clinical parameters on the gene expression at baseline and to evaluate if baseline gene expression might be a possible predictor of DAS28 development, we performed a Spearman correlation analysis. However, we only obtained weak to medium correlations (Supplementary Fig Figure 1C).

### FA transporter expression on CD8 from RA patients under different therapies

The FA transporters CD36 and GPR84 (Supplementary Figure 2A), which transport long-chain and medium-chain FAs [18,19], respectively, were the genes for which expression was the most effective for separating the baseline and 6-month time points for the subgroup of RA patients who received any therapy upon study enrolment. Together with FABP4, they are the main FA transporters in T cells (reviewed in [20]). Furthermore, we had previously observed that upon TCR-mediated stimulation, CD8, particularly from RA patients, imported considerable amounts of FA from the cell culture medium [4]. We therefore analyzed the expression of these FA transporters at the gene and protein levels on the surface of total CD8 and their functional subsets for healthy donors and RA patients under different therapeutic strategies.

At the gene expression level, *CD36* expression on the total CD8 pool did not differ between CNT and RA and was not influenced by the type of therapy, while *FABP4* and *GPR84* displayed a significant upregulation and downregulation, respectively, in RA DMARD-treated CD8 vs. CNT (Figure 2A). When subdividing the CD8 pool into its functional subsets, we did not observe any therapy-related changes in the gene expression of *CD36, FABP4*, and *GPR84* within each subset (Supplementary Figure 2B).

**Figure 2:**
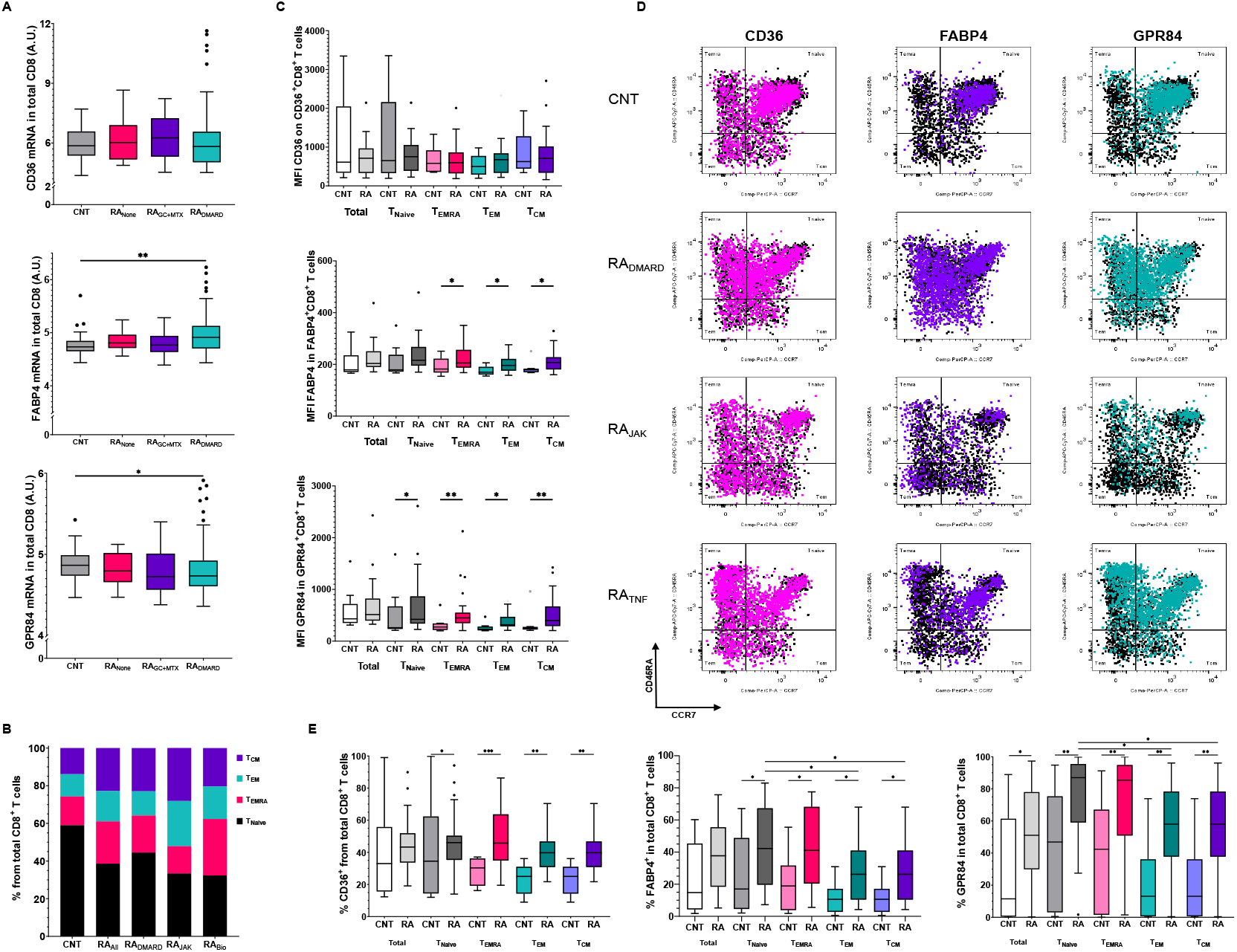
FA transporter expression on CD8 functional subsets from RA and CNT. The expression of CD36, FABP4, and GPR84 were analyzed on total CD8 and their functional subsets from RA patients and CNT. **A** Gene expression data from the Gene Omnibus datasets GSE97475, GSE97948, and GSE97810 with CD8 from RA patients according to therapy (CNT n=32; RA_no_ n=18; RA_DMARD_ n=149; RA_GC+MTX_ n=13). **B** Distribution of the CD8 functional subsets in CNT and RA under the different therapies. C-E FACS analysis of the FA transporters on CD8 functional subsets – **C** MFI values, **D** Representative dot-plot overlays of the FA transporter expressing population on the CD45RA CCR7 distribution in CD8 from CNT and RA patients sectioned according to the therapy strategies, **E** % of total CD8. For B CNT n=9, RA_ALL_ n=26, RA_DMARD_ n=13, RA_JAK_ n=4, and RA_BIO_ n=9. For **C**-**E** CNT n=9, RA n=26. NONE, no therapy; GC+MTX, glucocorticoids and Methotrexate; DMARD, common synthetic disease modifying anti-rheumatic drugs; JAK, JAK inhibitor therapy; BIO, bDMARD; TNF, anti-TNF therapy; T_NAIVE_, CD8 naïve subset; T_EMRA_, CD8 effector subset; T_EM_, CD8 effector memory subset; T_CM_, CD8 central memory subset.

Next, we analyzed the influence of different therapies on the CD8 functional subset distribution (Figure 2B). RA patients overall displayed a change in the distribution of the functional CD8 subsets when compared to CNT, while RA patients under JAK inhibitor (JAK) treatment and biologics therapy (BIO) presented significantly different subset distributions (CNT vs. RA _ALL_ X^2^=8.2, p=0.043; RA _JAK_ vs. RA _BIO_ X^2^=8.4, p=0.0391).

FABP4 and GPR84 median fluorescence intensities (MFI) were elevated in CD8 from RA in all functional subsets, but not in total CD8 (Figure 2C). The frequency of FA transporter populations within each functional subset was increased in RA compared to CNT for all three transporters, however it was less pronounced in the naïve compartment (Figure 2D-F). Therapy did not change the frequency of CD36 and FABP4 expressing CD8, however, GPR84 was lower in RA patients under JAK when compared to those receiving DMARD (Supplementary Figure 2C).

These findings could also be partially reproduced in CD8 from psoriatic arthritis (PsA) and spondylarthritis (SpA) patients compared to CNT (Supplementary Figure 3).

No correlations were found between the expression levels or frequency of FA transporter-expressing CD8 and clinical or demographic parameters (Supplementary Tables ST1-ST3).

### Metabolic profiling of patients with autoimmune arthritis

In order to evaluate the metabolic activity of the cells, we incubated CD8 in RPMI medium containing [U-^13^C]glucose either with or without TCR-mediated stimulation. Confirming our previous findings [4], CD8 from RA patients expressed an increased glucose uptake and lactate production at baseline as well as upon stimulation. CD8 from RA_BIO_ patients had higher fold-changes upon stimulation than CNT and RA_DMARD_ (glucose: CNT vs. RA_BIO_ p=0.0454, RA_DMARD_ vs. RA_BIO_ p = 0.0114; lactate: CNT vs. RA_BIO_ p=0.0363, RA_DMARD_ vs. RA_BIO_ p=0.0172) (Figure 3A and B). Also, FA uptake (Bodipy C16) was upregulated in RA CD8, however, it was not influenced by therapies or by other clinical or demographic parameters (Figure 3C, Supplementary Table ST4). The intracellular concentrations of palmitate (16:0), stearate (18:0), and arachidonate (24:4) did not significantly differ between the groups and upon in vitro stimulation but were tending to be increased in all disease groups (Figure 3D). The concentrations of unsaturated FA oleate (18:1), however, was elevated in CD8 from SpA patients compared to CNT, RA, and PsA CD8 (Figure 3D). Since FA-free serum was not used in the culture media, we were unable to quantify the percentage of FA derived from [U-^13^C]glucose.

**Figure 3:**
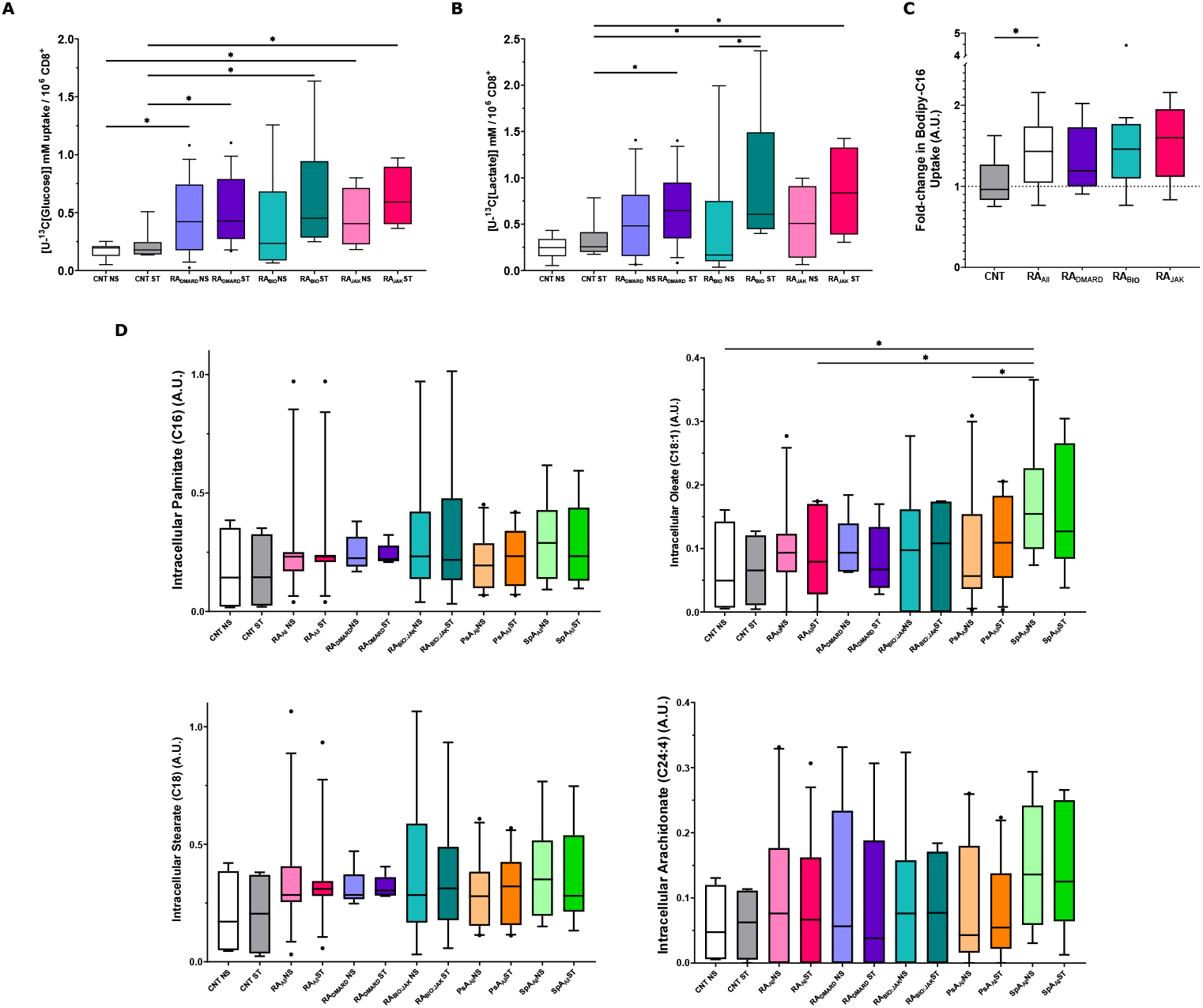
Metabolic analysis of CD8 from CNT and RA patients under different therapies. Total CD8 were cultured in vitro unstimulated (NS) or under TCR-mediated stimulation (ST). **A** Cellular [U-^13^C]glucose uptake. **B** Concentration of [U-^13^C]lactate in the cell culture supernatant. **C** FACS analysis of intracellular Bodipy-C16 uptake. CNT n=9, RA_DMARD_ n=13, RA_BIO_ n=9, and RA_JAK_ n=4. D MS analysis of intracellular FA concentrations of palmitate, oleate, stearate, and arachidonate (CNT n=4, RA_ALL_ n=11, RA_DMARD_ n=5, RA_BIO:JAK_ n=6, PsA_ALL_ n=10, and SpA_ALL_ n=9).

### Altered CD8 immune functions after in vitro inhibition of FA metabolism

To further investigate the FA metabolism in RA and the impact of the three transporters on CD8 functions, we used in vitro the following inhibitors: Etomoxir (FAO inhibition), TOFA (FAS inhibition), sulfo-succinimidyl oleate sodium (CD36i), CAS00657-03-8 (FABP4i), and GPR84 antagonist 8 (GPR84i). None of the inhibitors were observed to affect cell viability (Figure 4A). RA CD8 accumulated more neutral lipids upon TCR-mediated stimulation than healthy donors, and while the presence of the different inhibitors during stimulation reduced lipid accumulation in healthy CD8, only CD36i, Etomoxir, and GPR84i were able to reduce the neutral lipid content of RA CD8 (Figure 4B). *In vitro* stimulation induced proliferation (quantified by Ki67 expression) and activation (quantified by CD69 expression) in RA CD8 to a significantly larger extent than CNT CD8. Their proliferative capacity and the activation potential, however, were slightly reduced in RA CD8 in the presence of Etomoxir, FABP4i, and GPR84i and in CNT CD8 in the presence of CD36i, GPR84i, and TOFA (Figure 4 C-F). The cytotoxic capacity of the cells was assessed by Granzyme B expression levels. Etomoxir reduced the Granzyme B-expressing CD8 population as well as the MFI in RA CD8. TOFA treatment also induced a decrease of Granzyme B MFI in RA but not in CNT. In contrast to RA CD8, the Granzyme B positive population in CNT CD8 was decreased by CD36i, Etomoxir, and GPR84i (Figure 4 G-I).

**Figure 4:**
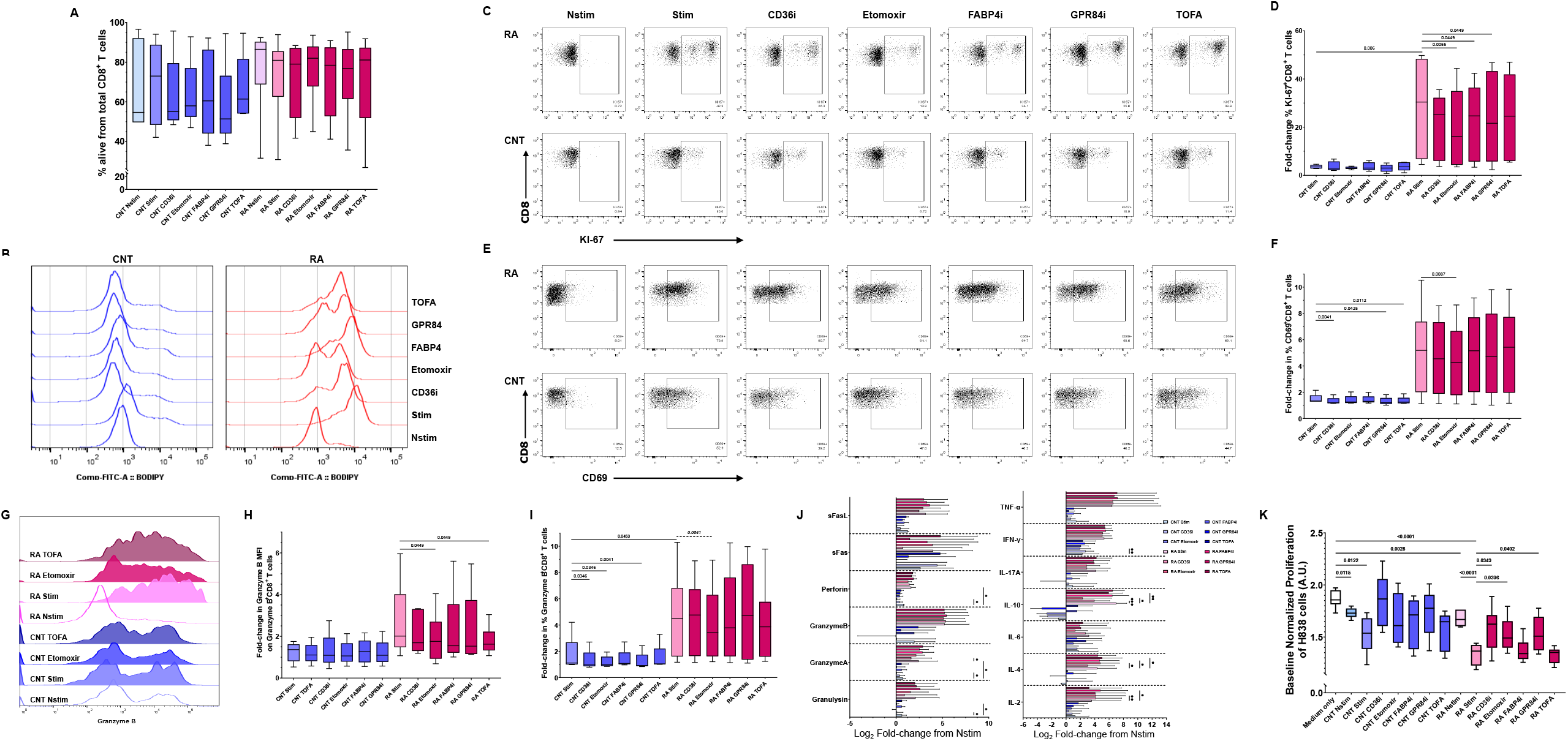
Effect of FA metabolism inhibitors on CD8 immuno-functional properties from CNT and RA patients. CD8 were cultured in vitro unstimulated (Nstim) or under TCR-mediated stimulation (Stim) and with the following inhibitors: Etomoxir (CPT1 inhibitor), CD36i (CD36 inhbitor), FABP4i (FABP4 inhibitor), GPR84i (GPR84 inhibitor), and TOFA (FAS inhibitor). Viability and effector functions were analyzed by FACS. **A** Cell viability was assessed using fixable viability staining. **B** FACS analysis of Bodipy (neutral lipids) uptake by total CD8 in the presence of the FA transporter inhibitors. **C**-**F** FACS analysis of the proliferation capacity (Ki67) and the activation (CD69) of CD8; fold-changes in comparison to Nstim. **G**-**I** FACS analysis of Granzyme B expression. Histogram display (**G**), fold-changes compared to Nstim in MFI (**H**) and % of total CD8 (**I**). J Analysis of cytokine release into the cell culture supernatant by a cytokine bead array. Displayed are fold-changes in concentrations compared to Nstim. **K** Assessment of the proliferation inhibition capacity of the secreted mediators in the supernatants on H383 cells (adenocarcinoma; non-small cell lung cancer) by Incucyte^®^. CNT n=5 and RA n=6.

For functional readouts, cytokine release by the cells was analyzed as well as the influence of secreted mediators present in the cell culture supernatants on the proliferation of the human lung adeno-carcinoma cell-line H838. As expected upon TCR-mediated stimulation, RA CD8 released more cytokines and cytotoxic mediators than controls (Figure 4J). In accordance with the above findings, CD36i, Etomoxir, and GPR84i affected CNT CD8 and decreased the secretion of cytotoxic and cytolytic molecules. Etomoxir, FABP4i, and GPR84i treatment resulted in a decrease in IL-10 secretion in RA CD8 and an increase in IL-4 and IL-2 secretion. H838 cell proliferation was significantly reduced upon addition of the conditioned media from stimulated CD8 from both groups (Figure 4K). In the presence of inhibitors H838 cell proliferation in conditioned media from CNT CD8 stimulated did not significantly change as compared to conditioned medium lacking inhibitors. However, H838 cells cultured in conditioned media from RA CD8 treated with CD36i, Etomoxir, or GPR84i proliferated significantly more than those cultured with conditioned media from TCR-stimulated CD8 without inhibitors. We could not find any statistically significant correlations between a reduction in the concentrations of cytolytic mediators and H838 proliferation (data not shown).

## Discussion

Research on the role of lipid metabolism in the maturation process of CD8 has shown that FAO is crucial for memory formation, while FA uptake from the extracellular milieu and FAS are essential steps in the metabolic rewiring of effector CD8 [12,21]. Studies on tumor-infiltrating CD8 have shown that hijacking CD8 lipid metabolism induced by the tumor microenvironment leads to a cytoplasmatic over-accumulation of oxidized lipids which undergo peroxidation and compromise the anti-tumoral effector functions [14]. However, while defects in lipid metabolism and how they contribute to inflammation have been thoroughly studied for RA CD4 [22], data on the role of lipid metabolism for the pro-inflammatory phenotype of RA CD8 is still largely missing. Additionally, it has been shown that RA patients treated with either MTX, the TNF-α blocker infliximab, or the JAK inhibitor tofacitinib present a significant increase in serological cholesterol levels and FAs [23–26], which suggests that such therapies might equally lead to alterations in lipid metabolism at the cellular level for CD8.

A recent large-scale, multiomics study on isolated total CD8 from RA patients presented a significantly different gene expression profile from healthy vaccinated controls [16]. Another gene expression analysis of sorted CD4 and CD8 functional subsets from small groups of RA patients under different therapies has revealed specific disease-related changes when compared to CNT [17]. Since none of these studies focused on lipid metabolism, we reanalyzed the two gene data sets focusing on the expression of genes directly related to lipid metabolism. The analysis of the data from the total CD8 set revealed that at the gene expression level, lipid metabolism appears to be differentially regulated in untreated early RA patients when compared to CNT. After these previously untreated early RA patients underwent six month of csDMARD therapy, predominantly with MTX, their CD8 lipid metabolism-related gene expression profile dramatically changed towards increased FA elongation, synthesis, transport, and oxidation. In general, a good clinical response to therapy with MTX (either alone or in combination with other DMARDs) was accompanied by a major change in the expression of FA metabolism-related genes and did not occur in patients who presented a good response to other types of DMARD therapies, while only a few genes were differentially expressed in the patients with no DAS28-CRP improvement after six months MTX therapy. Thus, an increase in FA metabolism in CD8 appears to be a characteristic specific for both, a good clinical response and to MTX. Actually, data from a serological study found that RA patients six months after MTX therapy initiation had higher concentrations of total cholesterol with both low- and high-density lipoprotein cholesterol increasing [27]. Another study in which MTX was administered to healthy rats reported an increase in hepatic lipid concentration but a decrease in serum lipid concentrations with a simultaneous increase in palmitoyl-CoA synthetase and carnitine palmitoyltransferase (CPT), supposedly due to reduced choline availability [28]. Together with our gene expression data, these studies suggest connectivity between the main MTX mechanisms of action in T cell-inhibition of adenosine release and impairment of JAK/STAT and NF-κB signalling (reviewed in [29])- and increased lipid metabolism in RA at systemic, tissue, and CD8 levels. Even though the number of samples from the CD8 functional subset analysis was limited, which partially restricted data interpretation, we could observe that in patients who did not receive treatment that the subsets with effector functions registered the largest number of total changes in lipid metabolism-related genes. It has been reported that altered FA metabolism can impair the anti-tumoral function of CD8 [29], and in CD4 impaired FAO inhibits the formation of the effector memory pool [29]. This suggests that the observed alterations in the expression of lipid metabolism-related genes in the effector subsets might fundamentally alter the effector functions of RA CD8 towards a chronic inflammatory response.

In contrast to memory CD8, effector counterparts express higher levels of the FA transporter CD36 and have increased intracellular palmitate levels [12]. All functional subsets of RA CD8 expressed CD36 more often, as well as FABP4 and GPR84, than CNT, and the cell surface expression of FABP4 and GPR84 were especially higher in RA effector and memory subsets. In mice, macrophages polarize towards a pro-inflammatory phenotype upon activation of the JAK/STAT-pathway by FABP4, while an agonist of GPR84 triggered TNF-α production in human peripheral blood mononuclear cells [30,31]. These studies may explain the decrease in the expression of these two transporters in RA patients receiving JAK inhibitor or TNF-blockade therapies.

Incorporating FAs into the metabolic portfolio might therefore help cells to survive in the nutrient deprived microenvironment of the synovium, akin to what has been seen in tumor-infiltrating cells. In those tumor cells, acetyl-CoA derived from imported palmitate was used as a carbon source for the TCA cycle to compensate for reduced glucose levels and to enable cells to maintain their anti-tumoral function [32]. Due to the limited sample size for some patients, we could only manage intracellular FA quantification for some samples. Nevertheless, we could observe that arthritis patients seem to have more intracellular palmitate (16:0), oleate (18:1), and stearate (18:0). Palmitate uptake induces T cell activation and increases their pro-inflammatory profile via PI3K/Akt signalling [32] and, in contrast to memory CD8, effector CD8 take up higher concentrations of palmitate [12]. Oleate and arachidonate (24:4) were shown to decrease proliferation and pro-inflammatory functions of Jurkat cells, though arachidonate was, however, already at a 10-fold lower concentration [33]. And whereas high intracellular concentrations of stearate triggered an inflammatory profile and apoptosis in mouse peritoneal macrophages, in mouse chondrocytes it promoted pro-inflammatory cytokine and lactate production and increased LDHA expression [34,35]. Therefore, it is plausible that in RA CD8 these FAs promote similar pro-inflammatory and metabolic profiles. Interestingly, the intracellular content of arachidonate, which is a precursor for the synthesis of systemically pro-inflammatory prostaglandins, did not appear to be affected in arthritis patients.

As FA-free serum culture medium was not used, we were unable to reconstruct whether these changes in intracellular FA concentrations were derived from increased FA uptake or FAS. Therefore, to analyze the impact of FA import from the extracellular milieu, we cultured CD8 in the presence of several FA transporter inhibitors. Upon TCR-mediated activation, CD8 switch to glycolysisin an mTORC1 dependent manner [36] and decrease OXPHOS, and in RA even naïve CD8 present such a metabolic profile [4]. Following activation of CD8 from arthritis patients, roughly 60% of ATP results from glycolysis [37] leaving the TCA cycle mainly for anabolic processes. Since aerobic glycolysis forces most of the pyruvate to be converted to lactate rather than to acetyl-CoA, the TCA cycle relies on acetyl-CoA resulting from FAO. The limiting step in FAO is the transport of acyl-CoA into the mitochondria by CPT1. Thus, without FA transport into the mitochondria, the already diminished mitochondrial acetyl-CoA pool in RA CD8 cannot be replenished. This may explain why RA CD8 exposed to the CPT1-inhibitor Etomoxir suffered a considerable loss of their immunofunctional capacity. In contrast, when FAS was inhibited, RA CD8 functionality was maintained. This suggests that FA uptake from the extracellular milieu might compensate for the reduced FAS as we did not see a major change in the total intracellular lipid content of TOFA-treated cells, nor on most immune functions. Even though we observed a dampening of RA CD8 immune activity upon inhibition of the FA transporters, single inhibition of each of them did not promote the same kind of functional alterations, which suggests that they can be compensated for by other FA transporters. Overall, our data show an increase in parameters related with FA synthesis and breakdown which suggests an increased FA turnover and a more dynamic FA metabolism. Based on these data and on those on FA-metabolism in RA CD4 [5], which are different from TCR-activated healthy T cells, one may consider the use of FAO and FA transport inhibition as new directions for future therapeutic strategies in RA, especially since the combined use of atorvastatin together with DMARD in a placebo-controlled study in RA patients has shown a significant improvement in DAS28 score [38]. Nonetheless, the potential hepatic toxicity of systemic FA metabolism inhibition requires that such interventions must first be tested in chronic polyarthritis mouse models or otherwise methods for cell-specific drug delivery be developed. Since some of these inhibitors are currently entering the clinical trial phase on cancer patients, this might expedite their use in RA.

## Data Availability

All data produced in the present study are available upon reasonable request to the authors. All data produced in the present work are contained in the manuscript.

## Acknowledgements

We thank Ms A. Funkert, all physicians of the Heidelberg University Hospital Rheumatology Outpatient Clinic and the Medical Clinic Blutentnahme team for coordinating and performing the blood collection. We are also very grateful to all of the patients and healthy volunteers who donated samples for the study.

## Funding

This work was supported by an internal grant from the Heidelberg Medical School to H-M Lorenz, and a grant from the German Research Foundation (Deutsche Forschungsgesellschaft) to M.M. Souto-Carneiro (SO 1402/1-1). F.V. Kraus was supported by an Add-On Fellowship of the Joachim Herz Foundation.

## Author contributions

Conceptualization: M.M.S.-C. and H.-M.L.; Methodology: M.M.S.-C., R.A.C., K.D.K, J.G., S.K., F.V.K., and H.M.L.; Formal Analysis: M.M.S.-C., R.A.C., K.D.K., S.K., and F.V.K.; Investigation: M.M.S.-C., K.D.K., S.K., and J.G.; Resources: H.-M.L., M.M.S.-C., K.D.K.; Writing-Original/ Draft: M.M.S.-C., F.V.K., and S.K.; Writing-Review & Editing: M.M.S.-C.., R.A.C., H.-M.L., K.D.K., S.K., F.V.K., and J.G.; Supervision: M.M.S.-C. and H.-M.L.; Funding Acquisition: M.M.S.-C. and H.-M.L.

## Competing interests

MMSC: Scientific support: Novartis, Pfizer.

HML: Scientific funding: Abbvie, Novartis, Pfizer, Roche; Consulting fees and honoraria: Abbvie, AstraZeneca, Actelion, Amgen, Bayer Vital, Boehringer Ingelheim, BMS, Celgene, GlaxoSmithKline (GSK), Galapagos, Janssen, Elli Lilly, Medac, MSD, Novartis, Pfizer, Roche, Sanofi, UCB; Travel support: Abbvie, AstraZeneca, Boehriner Ingelheim, BMS, Celgene, GSK, Gilead, Janssen, Elli Lilly, MSD, Novartis, Pfizer, Roche, Sanofi, UCB; Data safety monitoring and/or advisory board: Abbvie, AstraZeneca, Amgen, Boehriner Ingelheim, BMS, Celgene, GSK, Gilead, Janssen, Elli Lilly, Medac, MSD, Novartis, Pfizer, Roche, Sanofi, UCB

## Notes

### Author Declarations

The study was approved by the Institutional Ethics Committee of Heidelberg University, and is registred under case numbers S-096/2016 and S-969/2020.

