## Supplemental Materials and Methods for "Reduction of pro-inflammatory effector functions through remodeling of fatty acid metabolism in CD8^+^ T-cells from Rheumatoid Arthritis patients"

**Patients, Materials and Methods**

**Patients**

Patient recruitment and blood sample collection took place at the Division of Rheumatology outpatient clinic in Heidelberg. 80 mL of heparinized peripheral blood per study participant were collected from 31 patients with RA, 21 patients with SpA, 26 patients with PsA, and 14 healthy donors after giving their informed consent. RA and PsA disease activity was defined as remission (DAS28-CRP<2.6), low disease activity (DAS28-CRP≥2.6<3.2), or high disease activity (DAS28-CRP>3.2). SpA disease activity was defined by the clinical BASDAI score: inactive (BASDAI<2.0) and active (BASDAI>3.5). Therapy responders after 6 months were defined by a drop in DAS28-CRP of more than 0.6 points or when they retained a DAS28-CRP <2.6 . Therapy non-responders after 6 months were defined by an increase in DAS 28-CRP of more than 0.6 points or when their DAS28-CRP remained >3.2. Prospective patients and healthy controls were excluded from the study if any of the following applied: 1) malignant neoplastic disease or treatment at any time point; 2) pregnancy; 3) active viral and/or bacterial infection; 4) vaccination within the past 4 weeks; 5) denial of consent for study inclusion and blood donation.

**CD8 isolation, *in vitro* stimulation, and cell culture**

Peripheral blood mononuclear cells were isolated by density gradient isolation and CD8 were purified by magnetic-bead negative selection (MojoSort^TM^ Human CD8^+^ T Cell Isolation Kit (Biolegend). CD8 were cultured *in vitro* for 72 h at 37 °C under 5 % CO_2_ and 21 % O_2_ in RPMI 1640 medium (Gibco) containing 5 mM [U-^13^C]glucose (SIGMA-Aldrich), 10% heat-inactivated FCS (GIBCO), 100 U/mL penicillin, and 100 ng/mL streptomycin (both from GIBCO), either in the absence (non-stimulated, Nstim) or presence (stimulated, Stim) of anti-human CD28 (clone 28.1, 1 µg/mL, Biolegend) and plate-bound anti-human CD3 (clone OKT3, 2.5 µg/mL, Biolegend).

To modulate CD8 lipid metabolism, cells from 5 RA and 5 CNT were cultured with the following inhibitors: Etomoxir CAS 828934-41-4, 75 µM (inhibitor of FAO); 5-tetradecyloxy-2-furonic acid, 1 µM (inhibitor of FAS); sulfo-succinimidyl oleate sodium, 100 µM (inhibitor of FA transporter CD36); CAS00657-03-8 Inhibitor, 1 µM (inhibitor of FA transporter FABP4); GPR84 antagonist 8, 10 µM (inhibitor of FA transporter GPR84).

**Flow cytometry analysis of FA transporters in total CD8**

CD8 were stained with monoclonal antibodies against CD3, CD8, CCR7 (CD197), and CD45RA. Fatty acid transporters were stained with primary antibodies against (FAT) CD36 (Biolegend), GPR84 (Bioss), and FABP4 (R&D Systems) with subsequent addition of the secondary goat anti-rabbit IgG Alexa488 antibody (Cell Signaling Technologies) and anti-goat PE antibodies (R&D Systems). For palmitate uptake analysis, CD8 were stained with BODIPY FL C16 (ThermoFisher Scientific) according to the manufacturer’s instructions. For the assays with the FA metabolism inhibitors, cells were stained with either the viability dye Zombie Violet (Biolegend), Bodipy 493/503, anti-CD3, and anti-CD8 to determine intracellular total neutral lipid content, or with Zombie Violet, anti-CD69, anti-Ki67, anti-Granzyme B, anti-CD3, and anti-CD8 to quantify proliferation, activation, and cytotoxic mediator production.

After calibration with CST beads, single fluorochrome-stained cells were used for PMT setup and instrument compensation. All samples were run on an LSR II flow cytometer (Becton Dickinson). Minimum event rates were set to 5,000 events within the CD3^+^CD8^+^ gate after doublet exclusion. Resulting data were quantified using the FlowJo™ v10.8 Software (BD Life Sciences). CD8 subsets were identified using combined expression of CD45RA and CCR7: naïve CD45RA^+^CCR7^+^, effector memory RA-positive CD45RA^+^CCR7^-^, effector memory CD45RA^-^CCR7_-_ and central memory CD45RA^-^CCR7^+^.

**NMR analysis of media**

^1^H NMR spectra acquisition of cell culture media were performed using a 600 MHz Bruker spectrometer equipped with a 5-mm indirect detection probe. Each spectrum consisted of 43k points defining a spectral width of 7.2 kHz. A 30° radiofrequency observation pulse and a total repetition time of 10 seconds were applied to ensure full relaxation of all proton nuclei in the samples. Spectral analyses were performed using NUTSpro^TM^ NMR software (Acorn NMR Inc., Livermore, CA, USA). Prior to Fourier transformation, each free induction decay (FID) was multiplied by a decaying exponential with a decay constant of 0.2 Hz. Metabolite levels were quantified by deconvolution of the ^1^H NMR spectra using the line-fitting sub-routine of NUTSpro^TM^, with sodium fumarate {10 mM, dissolved in a 0.2 M phosphate buffer solution prepared using D_2_O (99.9%)} as an internal standard. Samples of cell-culture media consisted of 140 µL of media plus 35 µL of fumarate standard. The resonances due to ^13^C-enriched metabolites – the ^13^C satellites of lactate and acetate derived from the metabolism of [U-^13^C]glucose – are well resolved and allow the calculation of their concentrations for the evaluation of glycolysis and oxidative metabolism.

**Gene expression data analysis**

67 FA metabolism-related genes were identified using the Reactome database [1]. Gene expression data generated from RNA microarrays for total blood CD8 T cells from RA patients at baseline and after 6 months and from vaccinated (Hepatitis B) CNT were retrieved already normalized from the Gene Expression Omnibus (GEO: www.ncbi.nlm.nih.gov/geo/) data series GSE97476 [2]. Raw gene expression data generated from RNA microarrays for blood CD8 T cell subsets from RA patients under different therapeutic regimens and CNT were retrieved from the Gene Expression Omnibus (GEO: www.ncbi.nlm.nih.gov/geo/) data series GSE118829 [3,4]. Prior to analysis, the data sets were manually curated to select only the 67 genes related to FA metabolism. Gene expression analysis was conducted using the NASQAR packages [5]: “Gene Count Merger” for conversion of gene ids to gene names, duplicate removal and addition of pseudo-counts; “Create Meta Table” to merge clinical/demographic data to the gene expression data; and “DESeq2 Shiny” for normalization and differential gene expression analysis between groups. In the case of multiple probe-sets for one gene, the one showing the maximum average signal across the samples was selected. The heatmaps and the PLS-DA analysis were performed and generated using the statistical analysis package of Metabolanalyst 5.0 [6]. The Benjamini-Hochberg method with a false discovery rate (FDR) threshold of 5% was applied to account for multiple comparisons.

**H838 proliferation assay on IncuCyte**

The human adenocarcinoma (non-small-cell lung cancer) epithelial cell line H838 was maintained in high-glucose DMEM containing 10% heat-inactivated FCS, 100 U/mL penicillin, and 100 ng/mL streptomycin (all from GIBCO). Using 10,000 cells/well in a 96-well plate, H838 cells were cultured at 37°C in 5% (v/v) CO_2_ in an Incucyte S3 instrument for 5 days in either 75% fresh DMEM medium and 25% CD8-conditioned RPMI medium from CD8 culture with the different *in vitro* inhibitors or 75% fresh DMEM medium and 25% RPMI containing 5% glucose . The instrument was configured to whole-well mode and phase images were acquired every 2 h. Image data were analyzed with the vendor-supplied Incucyte S3 software (Incucyte version 2019B) using the Basic Analysis module to extract confluence data from the phase channel (analysis mask parameters available upon request). Data were corrected to zero time.

**Cytometric bead array for cytokines**

The titers of cytokines (IL-10, IFN-γ, IL-6, TNF-α, IL-2, and IL-17A) and cytotoxic molecules (Granzyme A, Granzyme B, Granulysin, soluble Fas, soluble Fas-Ligand, and Perforin 1) present in the media of CD8 cultures were determined using the NK/CD8 LEGENDPlex assay panel (Biolegend) according to the manufacturer’s instructions. CBA raw data analysis was performed using the cloud based LEGENDplex™ Data Analysis Software Suite (Biolegend) and concentrations were normalized and reported for a million cells.

**Mass spectrometry analysis of *de novo* fatty acid synthesis**

**FA methyl ester standards and identification means**

Methyl pentadecanoate (15:0), methyl palmitate (16:0), methyl oleate (18:1), methyl stearate (18:0), and a selection of other methyl ester FAs were purchased from Sigma. The FAs present in the CD8 samples were identified by a comparison of retention times to the standards together with confirmation by the MS library search function within the vendor-supplied data analysis software (MSD ChemStation G170EA E.02.02.1431). Methyl pentadecanoate, which was not present in CD8, was used as an internal standard for quantitation and was included at the methylation step as part of the BF3–methanol stock reagent during the preparation of the FA methyl ester samples for GC-MS analysis.

**Lipid isolation from CD8 culture and preparation of FA methyl esters for GC-MS analysis**

Total lipids were isolated from *in vitro* cultured CD8 by standard Folch extraction using chloroform-methanol (2:1). Transesterification of the lipids to FA methyl esters was accomplished using methanolic boron triﬂuoride as per literature [7], but briefly: BF3–methanol (1 mL, 14%, Sigma-Aldrich) was added to the CD8 chloroform extract, the vial tightly stoppered and incubated in a water bath at 90 °C for 4 h with vortexing for 5 s at hourly intervals. The reaction was quenched by cooling the reaction mixture to room temperature followed by the addition of H_2_O (0.5 mL). After raising the pH to 9–11 with aqueous NaOH (37%), chloroform (1 mL) was added and the two phases thoroughly mixed for 1 min followed by centrifugation for 30 s at 900 rpm after which the organic phase was removed using a Pasteur pipette and the solution filtered through a small wad of tissue in another Pasteur pipette on its way to a Mini-Vial. The filtrate was then taken to dryness using an air stream after which the FA methyl esters were taken up in 100 µL of CH_2_Cl_2_ and analyzed by GC-MS.

**GC-MS analysis of FA methyl esters**

Sample aliquots of the FA methyl esters were injected splitless (4 µL) into a 7890A A.01.13 GC system equipped with a 5975C inert MSD (Agilent Technologies) and ﬁtted with a HPB-5MS (5% phenyl methyl siloxane) column (30 m × 250 μm × 0.25 µm). Analyses were acquired with the following settings: inlet temperature, 250 °C; ﬂow rate, 1 mL/min helium; transfer line 150 °C; MS quadrupole, 150 °C; MS source, 230 °C; oven set at 40 °C initially, held for 3 min, then ramped to 300 °C at a rate of 16 °C/min, and finally held at 300 °C for 5 min (24.25 min total run time). Quantiﬁcation was performed using the vendor-supplied data analysis software (MSD ChemStation G170EA E.02.02.1431).

**Statistical analysis**

Continuous data variables were tested for normal distribution with a D’Agostino–Pearson omnibus normality test. Significant outliers were identified for each data set using the Grubb’s test/extreme studentized deviate test with an alpha value of 0.05 and those above the critical z-value were removed from the subsequent statistical analysis. Normally distributed unpaired data sets were analyzed using a one-way ANOVA followed by Fisher’s LSD test to determine individual p-values and a Brown–Forsythe test to determine the homogeneity of variances between groups. Normally distributed paired data sets were compared using a one-way paired ANOVA followed by a Greenhouse–Geisser correction for multiple comparisons between single groups. Unpaired data sets not following a normal distribution were compared between groups using a Mann-Whitney test followed by a Dunn’s multiple comparison test to determine the adjusted p-value. Within each group, the differences between Nstim vs. Stim vs. inhibitor-treated cells were assessed by a Wilcoxon matched-pairs signed rank test. Correlations between normally distributed data sets were determined using the parametric Pearson correlation coefficient. The Benjamini-Hochberg method with a false discovery rate (FDR) threshold of 5% was applied to account for multiple comparisons. Differences were considered statistically different for p<0.05.

**Supplementary Figure Legends SF1 – SF3:**

**Supplemental Figure 1: Gene and protein expression of lipid metabolism related genes**

**A** Heatmap of gene expression levels prior to therapy initiation in total CD8 from RA patients. **B** VIP scores plot for the most important genes for the separation of the gene expression at baseline and after 6 months of therapy in total CD8 from RA patients identified by the PLS-DA analysis. **C** Spearman correlation of clinical parameters for the RA patients and the lipid metabolism-related genes. Color coding represents Spearman’s R from -0.5 to 0.5.

**Supplemental Figure 2: and FA transporters in total CD8 and functional subsets**

**A** Gene expression of the FA transporters CD36, FABP4, and GPR84 in the CD8 functional subsets from CNT and RA patients under respective therapies (n=7-10 per subset per group). **B** FACS analysis of CD36, FABP4, and GPR84 expression in unstimulated CD8 from RA patients receiving respective therapies (RA_DMARD_ n=13, RA_JAK_ n=4; RA_Bio/TNF_ n=9).
Legend: no, no therapy; MTX, Methotrexate; DMARD, disease modifying anti-rheumatic drugs; JAK, JAK inhibitor therapy; TNF, anti TNF therapy; IL-6R, anti-IL-6 receptor therapy; Bio/TNF, anti TNF therapy and other bDMARD; T_N_, CD8 naïve subset; T_EMRA_, CD8 effector subset; T_EM_, CD8 effector memory subset; T_CM_, CD8 central memory subset.

**Supplemental Figure 3: FA transporter analysis on PsA and SpA patients**

FACS analysis of CD36, FABP4, and GPR84 expression on total CD8 and their functional subsets in CNT (n=9), PsA (n=25), and SpA (n=22) patients. **A** % of total CD8 FA transporter-expressing cells. **B** MFIs of the FA transporters in the functional subsets of CD8.
Legend: T_NAIVE_, CD8 naïve subset; T_EMRA_, CD8 effector subset; T_EM_, CD8 effector memory subset; T_CM_, CD8 central memory subset.

**Supplementary Tables ST1 – ST4**

**Supplementary Table ST1:** Correlation between **NSTIM CD36 MFI** levels and demographic and clinical parameters in RA patients and CNT (Spearman rank correlation coefficient and p-value)

| Group | RA | RA | CNT | CNT |
| --- | --- | --- | --- | --- |
| Parameter | **Correlation Coefficient** | **p-value** | **Correlation Coefficient** | **p-value** |
| Patient age |  | n.s. |  | n.s. |
| Patient BMI |  | n.s. |  | n.s. |
| Disease duration |  | n.s. | N.a. | N.a. |
| DAS28 |  | n.s. | N.a. | N.a. |
| Rheumatoid factor titer |  | n.s. | N.a. | N.a. |
| Anti-CCP^a^ titer |  | n.s. | N.a. | N.a. |
| CRP^b^ titer |  | n.s. | N.a. | N.a. |
| GFR |  | n.s. | N.a. | N.a. |
| Creatinine |  | n.s. | N.a. | N.a. |
| ^13^C-Lactate/10^6^ cells |  | n.s. |  | n.s. |
| Beta-blocker |  | n.s. | N.a. | N.a. |
| Glucocorticoid |  | n.s. | N.a. | N.a. |
| Methotrexate |  | n.s. | N.a. | N.a. |
| Leflunomid |  | n.s. | N.a. | N.a. |
| JAK-inhibitor |  | n.s. | N.a. | N.a. |
| TNF-alpha |  | n.s. | N.a. | N.a. |
| Other Biologics |  | N.a. | N.a. | N.a. |
| Parameter | **MFI of CD36  (mean ±** **SEM)** | **p-value** | **MFI of CD36  (mean ±** **SEM)** | **p-value** |
| Gender |  |  |  |  |
| Female | 785.1 ± 90.0 | 0.9961^c^ | 416.8 ± 113.0 | 0.4896^c^ |
| Male | 667.1 ± 167.6 |  | 1684.4 ± 607.6 |  |
| Biologics |  |  |  |  |
| No | 851.62 ± 92.9 | > 0.999^c^ | N.a. | N.a. |
| Yes | 934.00 ± 195.1 |  | N.a. | N.a. |

Legend: MFI, Median fluorescence intensity; N.a., not available; n.s., not significant; SEM, standard error of the mean; DAS28, Disease Activity Score; BASDAI, Bath Ankylosing Spondylitis Disease Activity Index; CRP, C-reactive protein; RF, rheumatoid factor; anti-CCP, anti-cyclic citrullinated peptide antibodies; GFR, glomerular filtration rate; MTX, methotrexate.

a) anti-CCP, antibodies against cyclic citrullinated peptides; b) CRP, C-reactive protein; c) p-values calculated by two-way ANOVA;

**Supplementary Table ST2:** Correlation between **NSTIM** **FABP4 MFI** levels and demographic and clinical parameters in RA patients and CNT (Spearman rank correlation coefficient and p-value)

| Group | RA | RA | CNT | CNT |
| --- | --- | --- | --- | --- |
| Parameter | **Correlation Coefficient** | **p-value** | **Correlation Coefficient** | **p-value** |
| Patient age |  | n.s. |  | n.s. |
| Patient BMI |  | n.s. |  | n.s. |
| Disease duration |  | n.s. | N.a. | N.a. |
| DAS28 |  | n.s. | N.a. | N.a. |
| Rheumatoid factor titer |  | n.s. | N.a. | N.a. |
| Anti-CCP^a^ titer | **-0.471** | **0.036** | N.a. | N.a. |
| CRP^b^ titer |  | n.s. | N.a. | N.a. |
| GFR |  | n.s. | N.a. | N.a. |
| Creatinine |  | n.s. | N.a. | N.a. |
| ^13^C-Lactate/10^6^ cells |  | n.s. |  | n.s. |
| Beta-blocker |  | n.s. | N.a. | N.a. |
| Glucocorticoid |  | n.s. | N.a. | N.a. |
| Methotrexate | **-0.486** | **0.041** | N.a. | N.a. |
| Leflunomid |  | n.s. | N.a. | N.a. |
| JAK-inhibitor |  | n.s. | N.a. | N.a. |
| TNF-alpha |  | n.s. | N.a. | N.a. |
| Other Biologics |  | N.a. | N.a. | N.a. |
| Parameter | **MFI of FABP4 (mean ±** **SEM**) | **p-value** | **MFI of FABP4  (mean ±** **SEM)** | **p-value** |
| Gender |  |  |  |  |
| Female | 228.4 ± 18.5 | > 0.999^c^ | 174.3 ± 5.5 | > 0.999^c^ |
| Male | 221.5 ± 9.1 |  | 229.6 ± 28.7 |  |
| Biologics |  |  |  |  |
| No | 442.69 ± 18.4 | > 0.999^c^ | N.a. | N.a. |
| Yes | 466.00 ± 24.2 |  | N.a. | N.a. |

Legend: MFI, Median fluorescence intensity; N.a., not available; n.s., not significant; SEM, standard error of the mean; DAS28, Disease Activity Score; BASDAI, Bath Ankylosing Spondylitis Disease Activity Index; CRP, C-reactive protein; RF, rheumatoid factor; anti-CCP, anti-cyclic citrullinated peptide antibodies; GFR, glomerular filtration rate; MTX, methotrexate.

a) anti-CCP, antibodies against cyclic citrullinated peptides; b) CRP, C-reactive protein; c) p-values calculated by two-way ANOVA;

**Supplementary Table ST3:** Correlation between **NSTIM** **GPR84 MFI** levels and demographic and clinical parameters in RA patients and CNT (Spearman rank correlation coefficient and p-value)

| Group | RA | RA | CNT | CNT |
| --- | --- | --- | --- | --- |
| Parameter | **Correlation Coefficient** | **p-value** | **Correlation Coefficient** | **p-value** |
| Patient age |  | n.s. |  | n.s. |
| Patient BMI |  | n.s. |  | n.s. |
| Disease duration |  | n.s. | N.a. | N.a. |
| DAS28 |  | n.s. | N.a. | N.a. |
| Rheumatoid factor titer |  | n.s. | N.a. | N.a. |
| Anti-CCP^a^ titer |  | n.s. | N.a. | N.a. |
| CRP^b^ titer |  | n.s. | N.a. | N.a. |
| GFR |  | n.s. | N.a. | N.a. |
| Creatinine |  | n.s. | N.a. | N.a. |
| ^13^C-Lactate/10^6^ cells |  | n.s. |  | n.s. |
| Beta-blocker |  | n.s. | N.a. | N.a. |
| Glucocorticoid |  | n.s. | N.a. | N.a. |
| Methotrexate |  | n.s. | N.a. | N.a. |
| Leflunomid |  | n.s. | N.a. | N.a. |
| JAK-inhibitor |  | n.s. | N.a. | N.a. |
| TNF-alpha  Other Biologics | N.a. | n.s.  N.a. | N.a.  N.a. | N.a.  N.a. |
| Parameter | **MFI of GPR84 (mean ±** **SEM)** | **p-value** | **MFI of GPR84  (mean ±** **SEM)** | **p-value** |
| Gender |  |  |  |  |
| Female | 747.1 ± 145.4 | 0.9877^c^ | 380.0 ± 32.7 | 0.9769^c^ |
| Male | 573.0 ± 81.3 |  | 751.8 ± 217.7 |  |
| Biologics |  |  |  |  |
| No | 964.15 ± 158.8 | > 0.999^c^ | N.a. | N.a. |
| Yes | 638.75 ± 40.2 |  | N.a. | N.a. |

Legend: N.a., not available; n.s., not significant; SEM, standard error of the mean; DAS28, Disease Activity Score; BASDAI, Bath Ankylosing Spondylitis Disease Activity Index; CRP, C-reactive protein; RF, rheumatoid factor; anti-CCP, anti-cyclic citrullinated peptide antibodies; GFR, glomerular filtration rate; MTX, methotrexate.

a) anti-CCP, antibodies against cyclic citrulinated peptides; b) CRP, C-reactive protein; c) p-values calculated by two-way ANOVA;

**Supplementary Table ST4:** Correlation between clinical or demographic data and transporter expression and Bodipy C16 – RA and CNT, Spearman R and p-value

| Group | RA | RA | CNT | CNT |
| --- | --- | --- | --- | --- |
| Parameter | **Correlation Coefficient** | **p-value** | **Correlation Coefficient** | **p-value** |
| Patient age |  | n.s. |  | n.s. |
| Patient BMI |  | n.s. |  | n.s. |
| Disease duration |  | n.s. | N.a. | N.a. |
| DAS28 |  | n.s. | N.a. | N.a. |
| Rheumatoid factor titer |  | n.s. | N.a. | N.a. |
| Anti-CCP^a^ titer |  | n.s. | N.a. | N.a. |
| CRP^b^ titer |  | n.s. | N.a. | N.a. |
| GFR |  | n.s. | N.a. | N.a. |
| Creatinine |  | n.s. | N.a. | N.a. |
| ^13^C-Lactate/10^6^ cells |  | n.s. |  | n.s. |
| Beta-blocker |  | n.s. | N.a. | N.a. |
| Glucocorticoid |  | n.s. | N.a. | N.a. |
| Methotrexate |  | n.s. | N.a. | N.a. |
| Leflunomid |  | n.s. | N.a. | N.a. |
| JAK-inhibitor |  | n.s. | N.a. | N.a. |
| TNF-alpha |  | n.s. | N.a. | N.a. |
| Other Biologics | N.a. | N.a. | N.a. | N.a. |
| Parameter | **MFI Bodipy C16  (mean ±** **SEM)** | **p-value** | **MFI Bodipy C16  (mean ±** **SEM)** | **p-value** |
| Gender |  |  |  |  |
| Female | 21363 ± 1365.2 | **>0.0001^c^** | 13758.3 ± 1064.7 | 0.7018^c^ |
| Male | 17759.1 ± 1585.7 |  | 14724.8 ± 2098.3 |  |
| Biologics |  |  |  |  |
| No | 17774.8 ± 1493.9 | 0.6324c | N.a. | N.a. |
| Yes | 21789.5 ± 1422.8 |  | N.a. | N.a. |

Legend: MFI, Median fluorescence intensity; N.a., not available; n.s., not significant; SEM, standard error of the mean; DAS28, Disease Activity Score; BASDAI, Bath Ankylosing Spondylitis Disease Activity Index; CRP, C-reactive protein; RF, rheumatoid factor; anti-CCP, anti-cyclic citrullinated peptide antibodies; GFR, glomerular filtration rate; MTX, methotrexate.

a) anti-CCP, antibodies against cyclic citrulinated peptides; b) CRP, C-reactive protein; c) p-values calculated by two-way ANOVA;
