## Supplementary figures and images for "Reduction of pro-inflammatory effector functions through remodeling of fatty acid metabolism in CD8^+^ T-cells from Rheumatoid Arthritis patients"

### Supplemental Figure 1

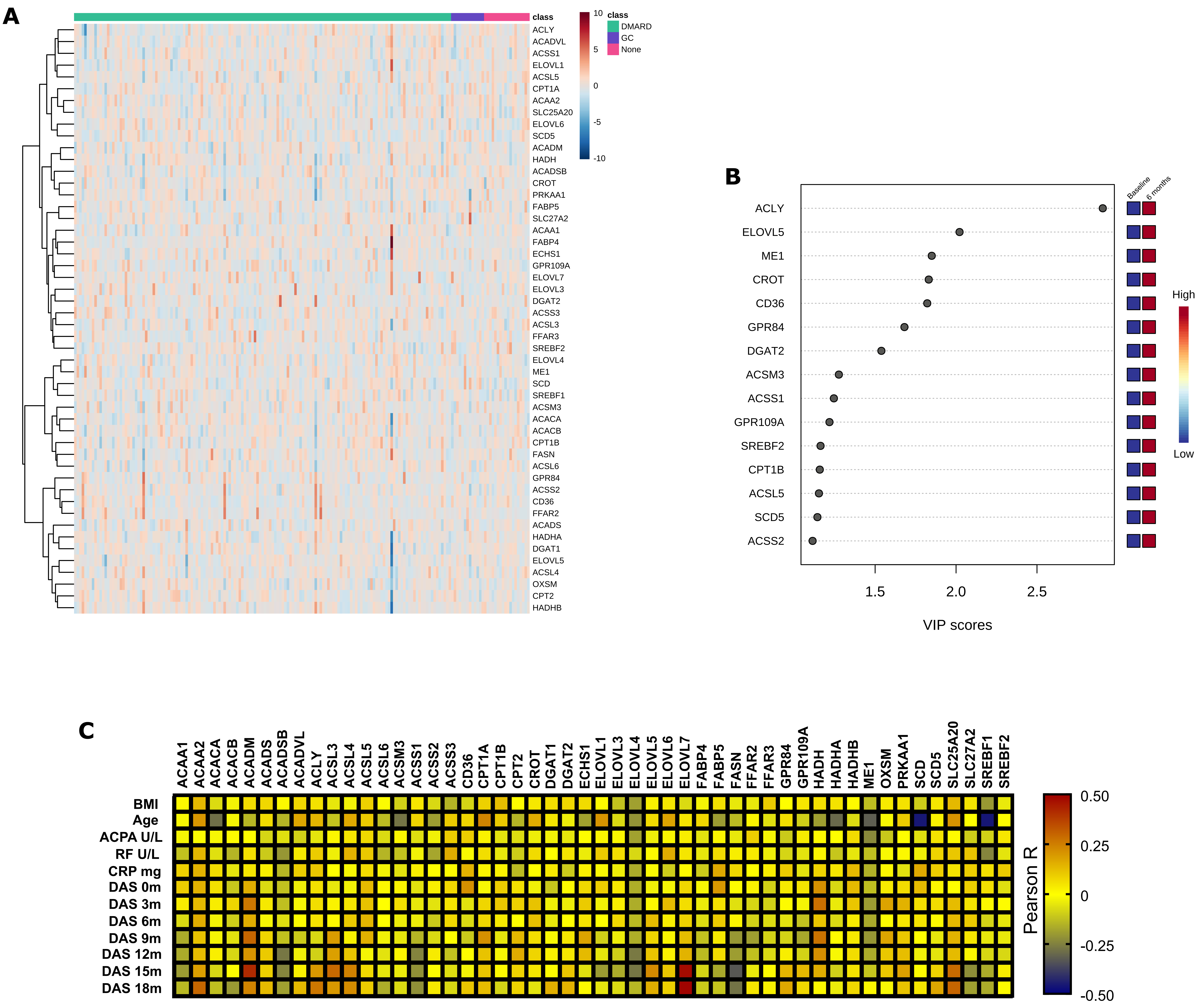

### Supplemental Figure 2

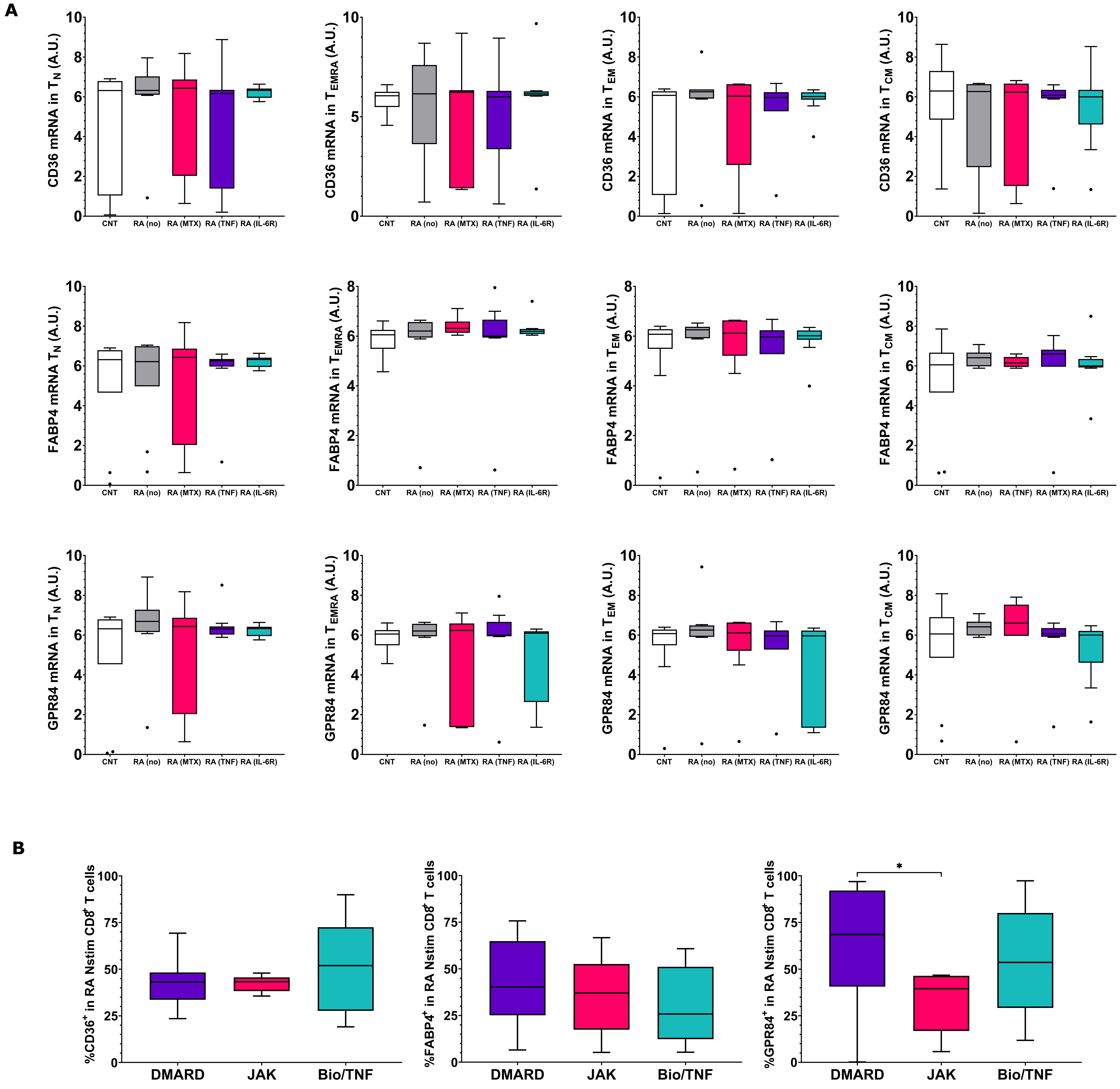

### Supplemental Figure 3

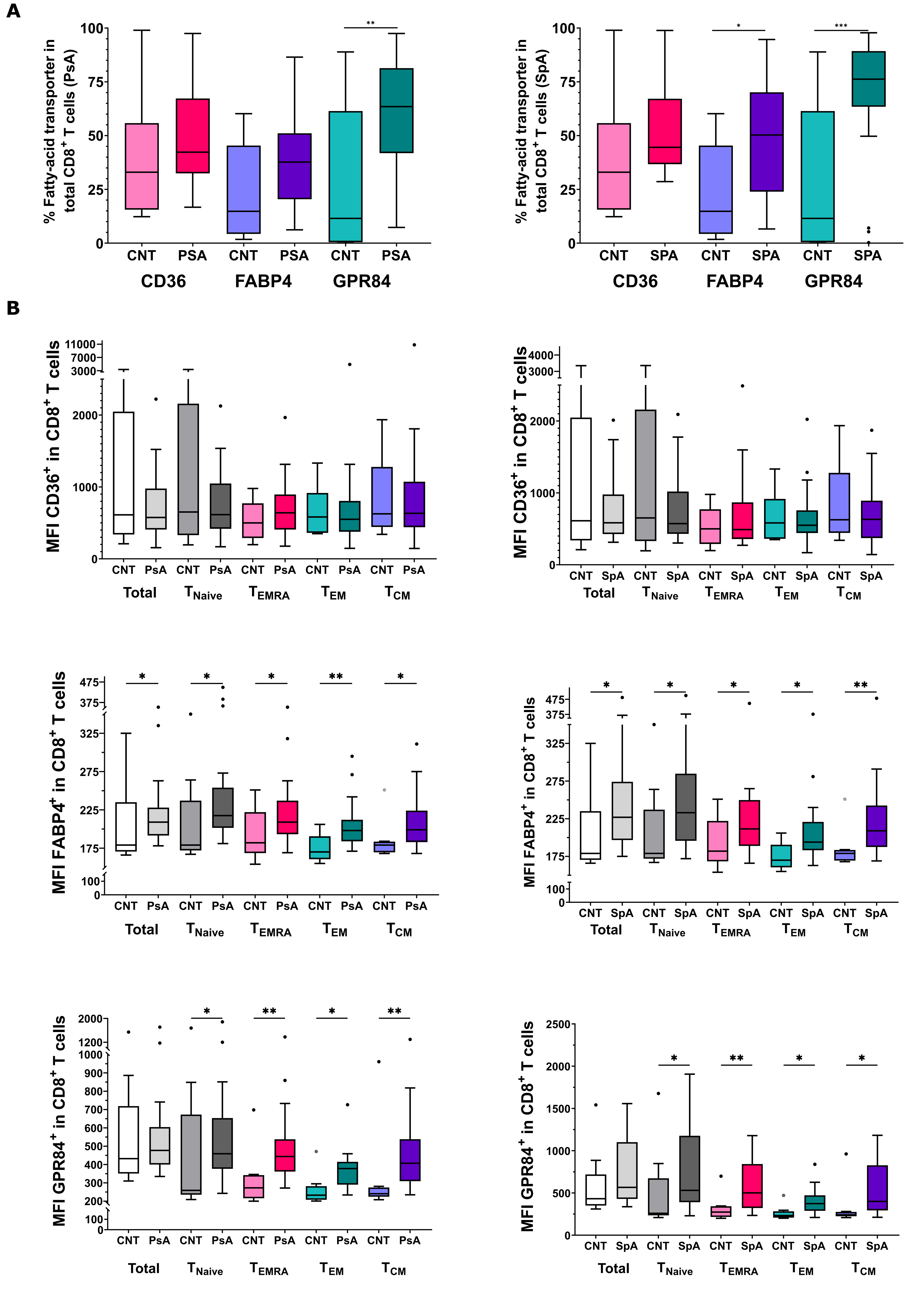
